# Effects on wrong-patient errors by limiting access to concurrently open ERH charts: A preliminary systematic mapping and synthesis review

**DOI:** 10.1101/2023.03.22.23287596

**Authors:** Lonn D. S. Myronuk

## Abstract

**Background:** Several recent outcome studies have been published looking at the effects of restricting electronic health record (EHR) user interfaces to limit the number of concurrently accessible patient records. Strong recommendations have been in place for several years to have user interfaces constrained to only display one patient chart at a time in order to reduce the risk of data (documentation, orders) being entered on the wrong patient (Joint Commission, 2015; ONC, 2016). This recommendation was made based on expert opinion rather than objective information, raising the question whether the accumulating evidence supports continued implementation of such chart access restrictions.

**Objectives:** This work reports a systematic mapping and synthesis review addressing research questions, “What is the evidence that restricting the number of concurrently open records reduces errors? (RQ1), “How effective is restriction of concurrently open charts at reducing wrong-patient errors? (RQ2), and “What additional inquiry is needed to make evidence-based policy decisions about restricting concurrent chart access? (RQ3).

**Methods:** A systematic search of CINAHL, PubMed, and Web of Science databases was performed with full search string specification to retrieve a result set that is the conjunction of result sets for concepts of *EHR, concurrently open charts*, and *medical error*. Of 407 studies identified and screened, five were eligible for inclusion in the qualitative synthesis review, and three were amenable to data extraction and pooled effect size calculation.

**Results:** None of the studies included for review found evidence of statistically significant change in wrong-patient error rates associated with implementing restriction in the number of patient records allowed to be open concurrently in the EHR. The combined OR for the pooled studies was 1.02 (95% CI 0.90 – 1.15) with low estimates for inter-study heterogeneity and no indication of publication bias.

**Conclusion:** There is no evidence that restricting the number of concurrently open records reduces errors (RQ1). It is not possible to definitively answer RQ2, but the magnitude of any yet to be detected beneficial effect that might be lost with lifting of chart access restriction can be no greater than an absolute risk increase of 33 errors per 100,000 ordering sessions. While it has been claimed that restricting the number of concurrently open EHR records is necessary for patient safety, the present review demonstrates that it is insufficient to attain a measurable improvement in error rates. Additional investigation of the usability and human factors aspects of EHR configuration decisions as well as knowledge of the impacts on clinical workflows will be necessary to provide policymakers, operational leaders, and practitioners with insight into the nature of the threats and opportunities with respect to safety, as well as the strengths and weaknesses of potential interventions.

---

The fundamental tenet of health care to “above all, do no harm” (Smith, 2005) is foundational to the triple aim of improved population health, enhanced patient care experience, and reduced per capita health care costs (Berwick et al., 2008). It is self-evident that harm accruing to patients as a result of health care will worsen their individual experiences, and that these harms will be cumulative to diminish across the population and increase the costs of care associated with the added morbidity. Extension of the framework for health care system imperatives to include provider work experience (quadruple aim, Bodenheimer & Sinsky, 2014) and health equity (quintuple aim, Itchhaporia, 2021) do not diminish the paramount importance of the non-maleficence principle to avoid causing harm, but expand the scope of what policy makers and operational leaders must consider when appraising risk of harm: Potential harms to the healthcare practitioner workforce must be taken into account, as must the differential effects of policy decisions across varied segments of society in the population as a whole. It is in this context that accreditation guidelines and recommendations for practice standards have arisen, such as the admonition to restrict the number of patient charts permitted to be concurrently open in an electronic health record to just one (Joint Commission, 2015; Office of the National Coordinator for Health Information Technology, 2016). This particular recommendation has been of some concern as it is based on expert opinion without supporting objective evidence (Adelman, 2019) and could be associated reduced provider workflow efficiency, increased fatigue, and unintended paradoxical increase in medical error (Su, 2019). The Columbia University group led by Adelman (2019) addressed the question of risk in a prospective randomized clinical trial looking at the effects of limiting the number of concurrently open EHR charts on the rate of erroneously entered patient orders, the first study of its kind. Other quasi-experimental studies have also been published, making the topic amenable to systematic review.

In earlier work, a proposed method was presented for mapping the extant health informatics literature on the effect of the number of concurrently openable charts has upon the rate of wrong-patient electronic health record (EHR) errors (Myronuk, 2022). Three research questions were identified:

**RQ1:** What is the evidence that restricting the number of patient charts able to be open concurrently in an electronic health record leads to a reduction in the rate of “wrong patient” data entry errors?

**RQ2:** How effective is restriction of the number of charts permitted to be open concurrently in an EHR at reducing the rate of “wrong patient” data entry errors?

**RQ3:** What additional inquiry (further reviews or primary research) is needed to inform specific organizational policy decisions regarding the optimal configuration setting for concurrent chart access?

This study attempts to address these research questions and elucidate the strength of relationship between the number of charts a provider may have open in the computer in front of them and the chance of them inadvertently entering information into a record that was not intended. While this is of immediate interest to those practitioners dealing with patients and their electronic records, all of whom aspire to first do no harm and provide safe care, it is of broader interest to stakeholders in the healthcare system. This includes patients for whom erroneous chart entry could result in adverse health consequences (including discomfort, disability, or death), and the stewards of the healthcare system – its policymakers and operational leaders – who are answerable for its efficient and equitable delivery of services within available resources. The principal goal of the present work is to provide stakeholders with knowledge to inform the decision-making around EHR configuration and use to best support the quintuple aim.

## Methods

The literature search used was that described in Myronuk (2022), which follows PRISMA-S structure (Rethlefsen et al., 2021). Individual searches were performed on CINAHL, PubMed, and Web of Science databases, utilizing the search strings listed in Table 1. Backward snowball sampling (Jalali & Wohlin, 2012) was utilized to add relevant publications to the review set. As the systematic mapping review does not entail research on human (or animal) subjects, no institutional review board approval was required (Sullivan, 2011).

**Table 1.** Searches in databases

| Database | Search |
| --- | --- |
| CINAHL<sup>1</sup> | (electronic health record* OR electronic medical record* OR emr OR ehr)<br>AND (concurrent* N2 open OR chart* N2 open OR record* N2 open) AND (medical errors OR patient harm OR wrong patient errors OR wrong patient) |
| PubMed | ("Electronic Health Records"[MeSH Terms] OR "electronic health record*" [All Fields] OR "EHR" [All Fields] OR "EMR" [All Fields]) AND (("concurrent*" [All Fields] AND "open" [All Fields]) OR ("open" [All Fields] AND "concurrent*" [All Fields]) OR (("chart*" [All Fields] AND "open" [All Fields]) OR "open chart*" [All Fields]) OR (("record*" [All Fields] AND "open" [All Fields]) OR "open record*" [All Fields])) AND ("Patient Harm" [All Fields] OR "Patient Harm" [MeSH Terms] OR ("medical errors" [MeSH Terms] OR ("medical" [All Fields] AND "errors" [All Fields]) OR "medical errors" [All Fields] OR ("medical" [All Fields] AND "error" [All Fields]) OR "medical error" [All Fields]) OR "wrong patient" [All Fields]) |
| Web of Science | (ALL=(electronic health record*) OR ALL=(EHR) OR ALL=(EMR)) AND<br>(ALL=(concurrent* open) OR ALL=(record* open) OR ALL=(chart* open)) AND<br>(ALL=(medical error*) OR ALL=(patient harm) OR ALL=(wrong patient)) |

<sup>1</sup> The Boolean search phrase for CINAHL had expanders applied (related words, full text search, equivalent subjects)

### Knowledge management tools

The toolchain for management of the literature search results, screening, data extraction, and analysis included *EndNote* (The EndNote Team, 2013), *Covidence* (*Covidence systematic review software*), *R* (R Core Team, 2022), *RStudio* (RStudio Team, 2022), and the associated software packages *tidyverse* (Wickham et al., 2019) and *meta* (Balduzzi et al., 2019). Citations retrieved from each source database were imported into *EndNote* where duplicates could be identified and removed. The list of unique citations was then uploaded into *Covidence* for stepwise screening of titles and abstracts, full text review, and data extraction. The extracted data were then ported into R using RStudio where statistical analyses were executed.

### Data Analysis

A total of 407 candidate studies were identified using the specified search strategy (Table 2). For inclusion, studies must have examined an implemented EHR in clinical use, where the subject EHR can be configured to allow more than one patient chart to be loaded concurrently in the user interface, and to have reported observed rates of medical errors in association with number of number of charts allowed open concurrently. Studies with paper-based and hybrid paper-electronic systems were excluded, as were studies of EHR performance in clinical simulation and training scenarios or pre-production settings. Studies that did not have available full text in English were excluded, and only peer-reviewed publications were excluded (no content from pre-print servers, or grey literature). Figure 1 shows the PRISMA flow diagram for screening and selection of studies for this review. A low threshold was employed for including a paper for full-text review, and any items that were of uncertain relevance after screening of title and abstract were brought forward to be read in full.

**Table 2.** Search results for source databases

| Database | Search results |
| --- | --- |
| CINAHL | 53 |
| PubMed | 114 |
| Web of Science | 240 |

**Figure 1.**
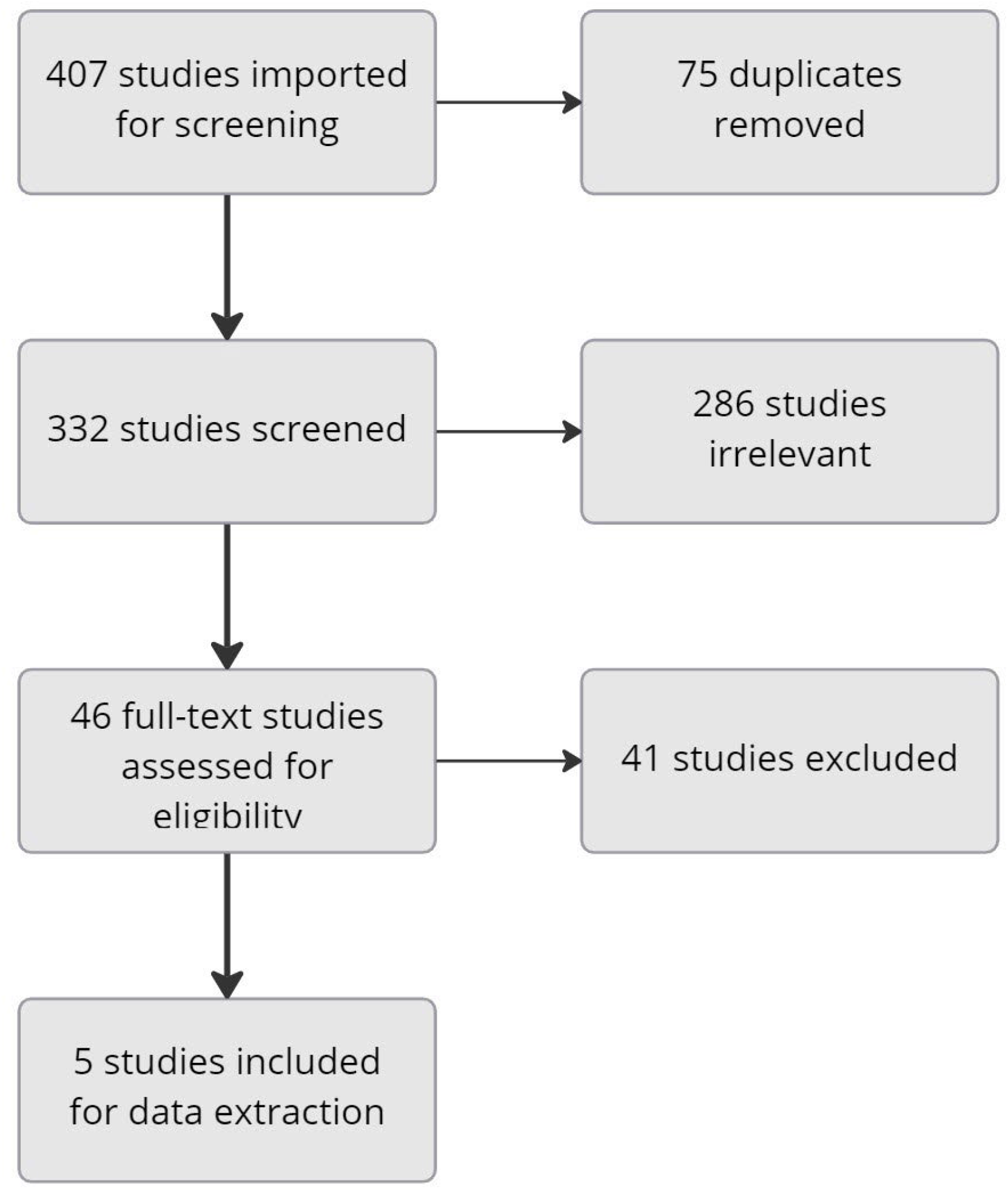
PRISMA Flow Diagram showing study identification and selection.

Five published reports met criteria for inclusion after full-text review (Table 3). All studies retained the null hypothesis that there was no difference in observed rate of wrong-patient errors between EHR configurations where number of concurrently open charts was unrestricted (up to 4 charts open) and configurations restricting multiple chart access. The study of Adelman and colleagues (2019) is a prospective randomized clinical trial, the remainder are retrospective time series cohort studies. The settings for the various studies varied, with emergency department and general inpatient settings most represented. All studies were carried out in organizations where a commercially available EHR had been implemented, three utilizing EpicCare and two using Cerner. The wrong-patient error events were defined in each study based on the Wrong-Patient Retract and Reorder (WP-RAR) Measure, which states an error event occurs when an order is placed on a patient within an EHR, is retracted within 10 minutes, and then the same clinician places the same order on a different patient within the next 10 minutes (National Quality Forum, 2016). As data were generated by query reports run in the subject EHRs, missing data was reported not to be an issue for of the studies.

**Table 3.**
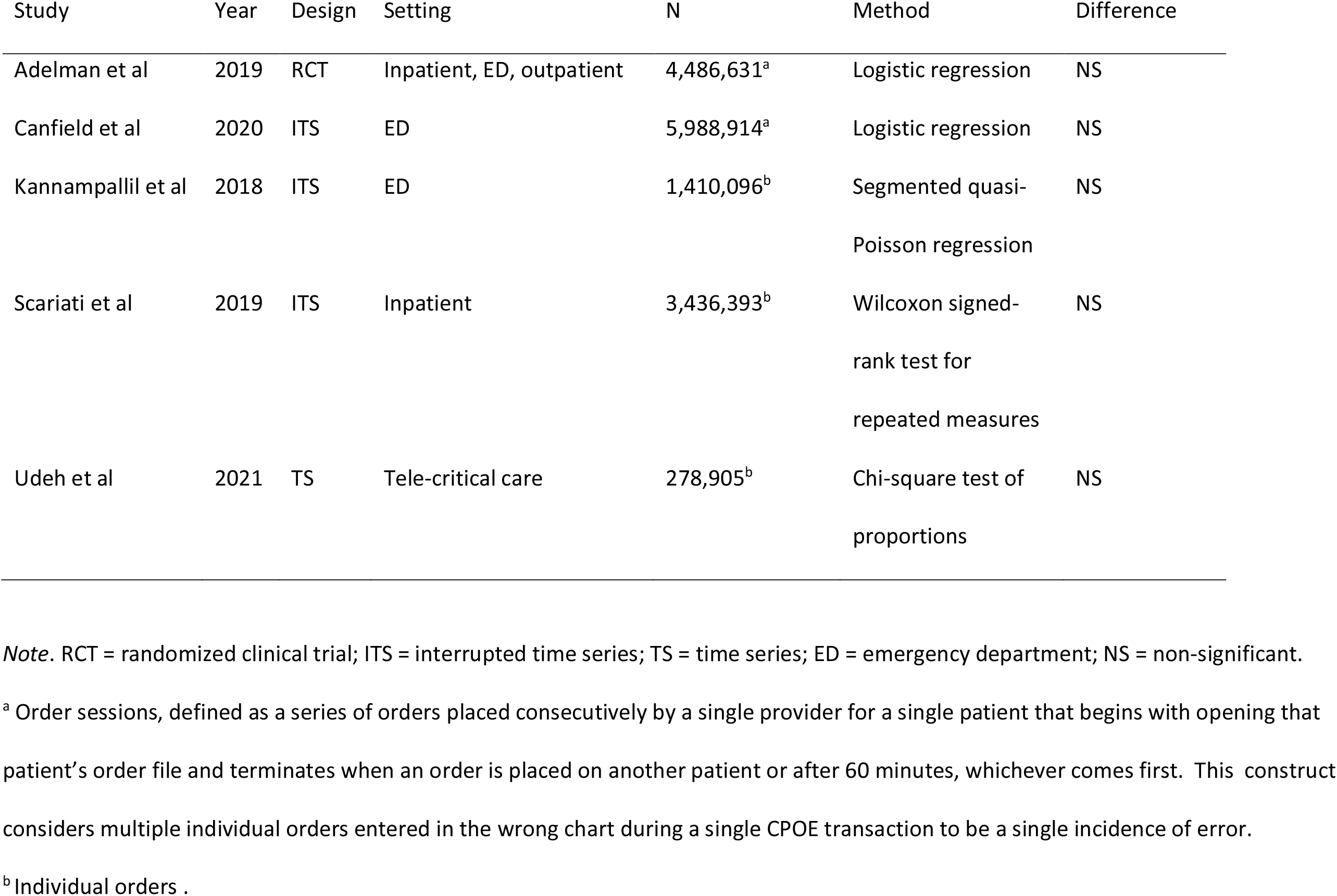
Studies meeting criteria for inclusion in review

The study of Canfield et al. (2020) reported error event rates per thousand order sessions for restricted and unrestricted conditions of 2.2/1000 and 2.4/1000, respectively. They reported the total number of order session instances across the entire study (*n* = 5,988,914), but did not give the subtotals for the two conditions and hence could not be included in computing a pooled effect size for all studies. Similarly, the paper by Scariati et al. (2019) did not provide sufficient detail regarding observed order volumes and error event counts across conditions to be included.

The outcomes (odds ratios) of remaining three studies were combined to calculate a pooled binary effect size (Harrer et al., 2021) using R (R Core Team, 2022; RStudio Team, 2022). A random effects model was assumed. Studies were assigned weighting with use of the Peto method (Yusuf et al., 1985), as it performs well when events are very rare (Bradburn et al., 2007). Heterogeneity between studies was assessed with the *I*^2^ Statistic (Higgins & Thompson, 2002), and the restricted maximum likelihood estimator was used to compute the heterogeneity variance *τ* ^*2*^ (Viechtbauer, 2005).

## Results

The results of the pooled analysis are presented in Figure 2 as a forest plot with accompanying tabular data values. The overall odds ratio for the random effects model is 1.02, 95% CI 0.90 – 1.15, which does not reject the null hypothesis of no difference in wrong-patient error rate between EHR configurations. Heterogeneity was low among the included studies, and the prediction interval (Inthout et al., 2016) straddles unity suggesting that future studies would fall in the range consistent with no intervention effect. A funnel plot (Figure 3) was generated to examine for reporting bias in the published studies. As no studies fall outside the area of statistical non-significance and there is symmetry around the computed average effect size, there is no imputation of publication bias among the included studies.

**Figure 2.**
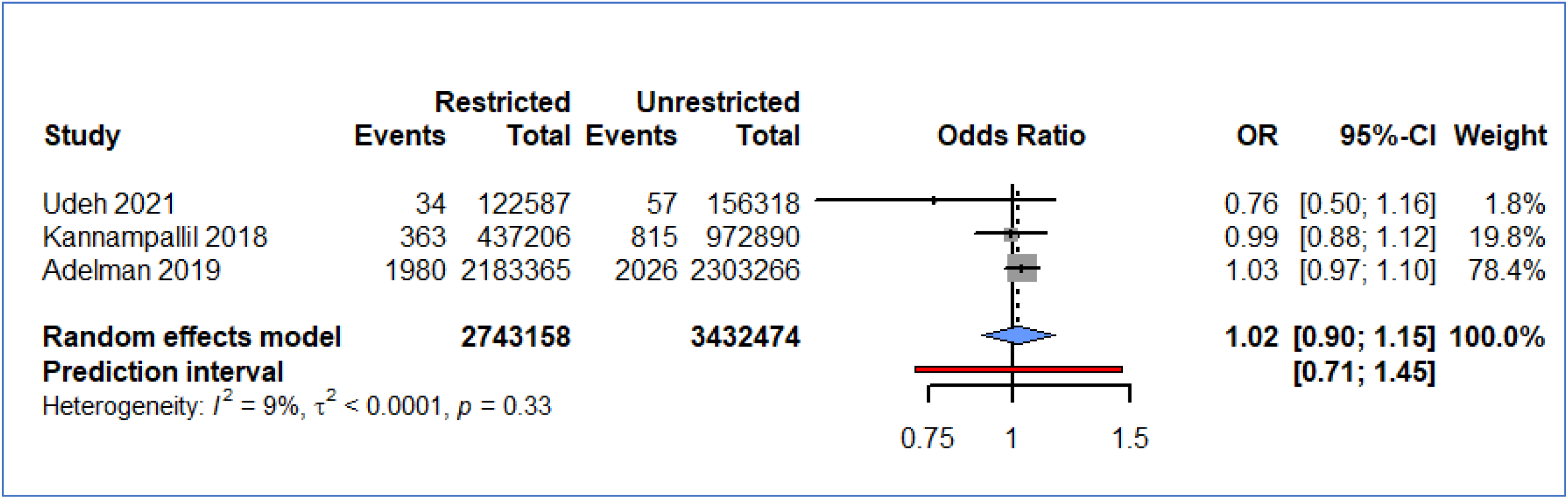
Pooled analysis results for effect of number of charts allowed open on rate of wrong-patient errors in the EHR. *Note*. Details on meta-analytical method: Peto method; Restricted maximum-likelihood estimator for τ^2^; Q-Profile method for confidence interval of τ^2^; Hartung-Knapp (HK) adjustment for random effects model (df = 2).

**Figure 3.**
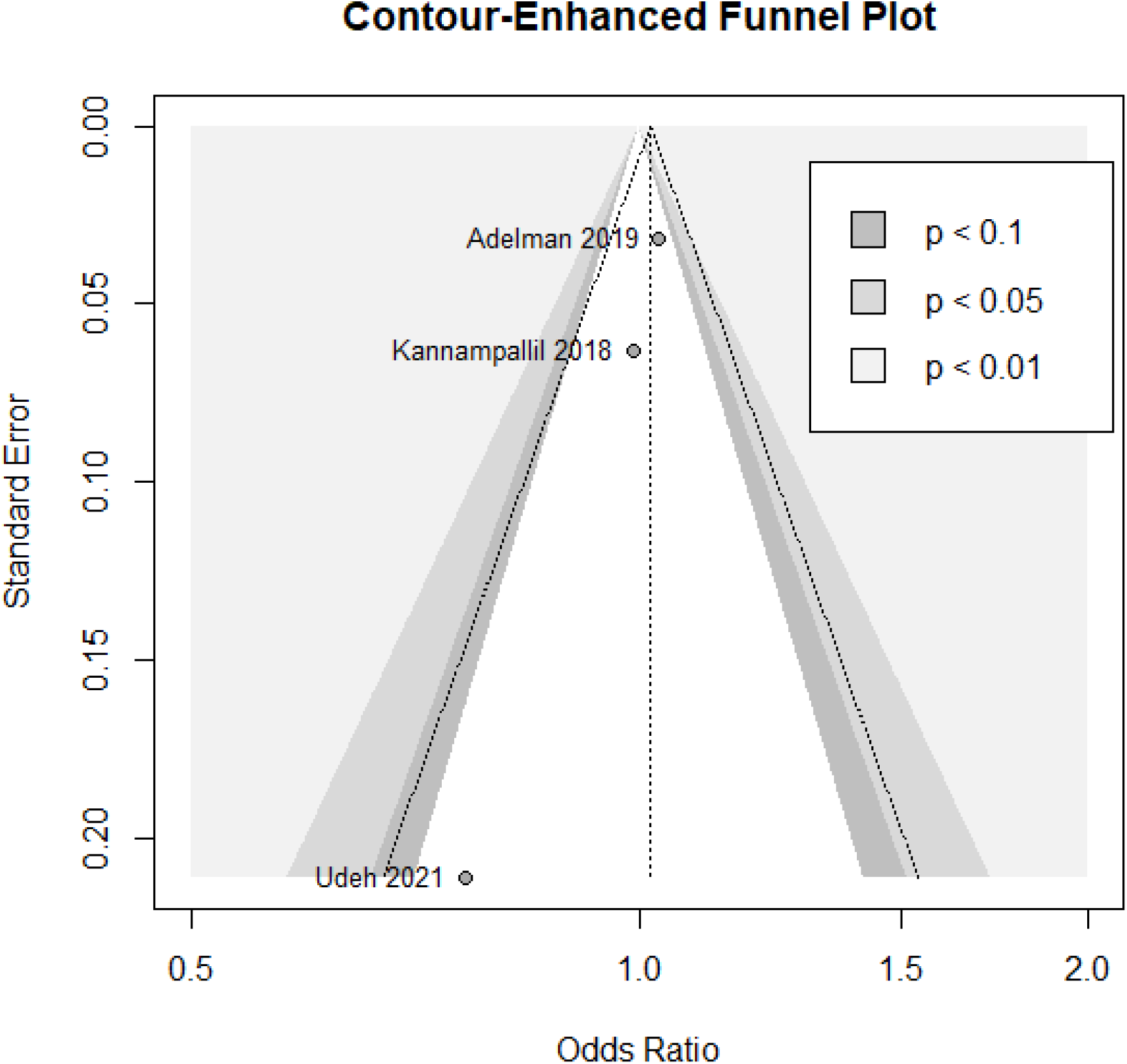
Funnel plot with pseudo 95% confidence limits

## Discussion

The results of the systematic mapping of the literature identified five independent studies, one randomized clinical trial and four retrospective time series cohort studies, all of which reported no statistically significant difference was found in wrong-patient error rates between conditions of restricted versus unrestricted access to multiple charts in the EHR. Of the five studies, three presented data that permitted pooled effects to be computed, the results of which are also a finding of no statistically significant difference in error rates between EHR configuration conditions. Analysis of the pooled study data demonstrated low heterogeneity between studies and did not suggest the presence of publication bias. Given the foregoing, the answer to RQ1 can be stated with reasonable confidence:

**RQ1:** There is currently no evidence that restricting the number of patient charts able to be open concurrently in an electronic health record leads to a reduction in the rate of “wrong patient” data entry errors.

It may be somewhat reassuring for proponents of unrestricted EHR configurations that no difference in error rates has been ascertained (Wachter et al., 2019), but one must ask whether this is sufficiently persuasive evidence upon which to base a decision to implement a change in organizational policy. In health care generally, before recommending an intervention we seek evidence to support the assertion that the intervention will have the desired outcome effects. Had no chart restrictions been in place, the results here would not be accepted as evidence to support the implementation of open-chart restrictions to improve patient safety, based on a type-I error rate of 5% (α ≤ 0.05). For organizations that have not (yet) implemented the JC/ONC recommendation to limit the number concurrently open charts to one, it is valuable to know that the available evidence does not support this recommendation as being effective at reducing wrong-patient errors in the EHR. It follows that any business case to be made for implementing the recommendation must be based on rationale other than anticipation of reduced medical error rates^1^.

The reality is that many organizations do have extant policies restricting the number of allowable concurrently open charts in the EHR (Adelman et al., 2017). In the face of national guidelines that unequivocally recommend implementing chart restrictions, policymakers need evidence to inform a related but different decision: What is the risk of committing type-II error and *falsely accepting* the null hypothesis of no difference between configurations? That is to say, what are the chances that there actually is a real-world difference in outcomes between EHR configuration setting states, but our measurements have not been able to discern it? Regulators and operational leaders will want to know what the chances are that *lifting restrictions* might reveal such an unmeasured effect. With respect to RQ2, the present study offers no illumination. We have not ascertained the presence of an intervention effect, but this is not synonymous with ascertaining the absence of any intervention effect.

The type-II error rate (β) describes the chance of accepting a null hypothesis (no difference between conditions) when it is incorrect, and an actual difference does exit. Beta error rates are related to the statistical concept of *power* (1-β) which is defined as the probability that the null hypothesis will be *correctly* rejected. By convention, a study with power of 0.8 is considered adequate for detection of true treatment effects (Harrer et al., 2021), but the resulting 20% type-II error rate may not be acceptable for our purposes: patient safety. In the context of health and safety, studies of toxicology exemplify the importance of being able to establish trust in the meaning of results where “no significant difference” is found. For studies of this kind, the confidence interval upper bound is used to establish the upper limit for an unknown (i.e., undetected) true effect (Mair et al., 2020). For the present study, this means that polled results indicate that the largest effect that unrestricted EHR chart access could have on wrong-patient error rates is an odds ratio of 1.15, and given the low base incidence of these types of EHR errors (<<10%) the odds ratio closely approximates the ratio of relative risk (Zhang & Yu, 1998). Hence, the relative risk of wrong-patient EHR errors with multiple concurrently open charts will not exceed 1.15. The absolute risk of these errors is low, based on the event rates reported in the reviewed studies, as well as the preceding work establishing validity of the WP-RAR measure (Adelman et al., 2013). Error rate values reported for unrestricted EHR configurations range from 28/100,000 to 220/100,000 in the reviewed studies, meaning that the maximum increase in absolute risk would be no more than 33 errors per 100,000 order events.

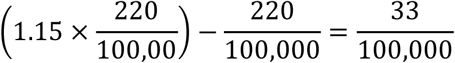

The policymakers and operational leaders responsible for healthcare outcomes will need to make judgements in their respective organizational contexts whether the potential loss of undetected beneficial effects of limiting concurrent chart access which could amount to this small absolute increase in error rate is justifiable in terms of improved workflow efficiency and diminished cognitive burden for providers.

### Future study (RQ3)

This systematic mapping and synthesis review points to the feasibility of a formal systematic review and meta analysis, where the quality of the primary research and risk of bias can be more rigorously examined. Within such a systematic review framework it would be possible to obtain data elements not included in published papers that would allow more studies to be included in the meta analysis than was possible in the computation of pooled effects reported here. This would further refine the precision of imputed effect size estimates.

Beyond the efforts to examine WP-RAR events as an objective outcome of the EHR configuration changes studied, it is clear when viewed through the lens of the quintuple aim of healthcare that there needs to be concurrent evaluation of outcomes for the healthcare system in domains of provider experience, cost, and health equity. Survey data has suggested that the restricted configuration is less acceptable to providers, and in some cases is associated with work-arounds that may increase patient risk (Southern et al., 2019). Further investigation of the usability and human factors aspects of the proposition to limit concurrent access to multiple patient records in the EHR is needed to bring understanding of the impact of such changes on speed and efficiency of care provision, as well as cognitive effort and provider fatigue. With such insights, it is possible to draw a more nuanced conclusion about the true cost-benefit ratio of a policy decision to limit concurrent chart access.

## Conclusion

The decision whether to restrict EHR affordance so as not to allow concurrent access to multiple patient records is contentious, with organizations being divided in their alignment to the JC/ONC recommendation of only permitting access to a single chart at a time (Adelman et al., 2017). When contemplating changes to complex sociotechnical systems it is valuable to reflect on the question, “What problem is it we are attempting to solve?” Not every safety measure is amenable to being tested in a randomized controlled trial, and it would be unethical to withhold safety measures with self-evident benefits solely for the sake of intellectual rigor — for example, parachutes when jumping from aircraft (Smith & Pell, 2003). It seems to stand from first principles that multiple concurrently open charts would entail a hazard for mistakenly entering data into an unintended patient chart, yet the evidence to date does not support this contention. Is it correct to conclude that the facility to open multiple charts concurrently is immaterial to the risk of wrong-patient errors, or is it that the potential benefits of a restricted EHR configuration are unable to be realized given other aspects of the complex business of health care delivery? Extending the parachute analogy, if we observe that the morbidity and mortality rate is the same when jumping from an aircraft with or without a parachute, the finding may be better accounted for by understanding the physics of bailing out at 150’ altitude whilst travelling 300 m.p.h. than by any consideration of parachute use!

## Data Availability

All data produced in the present study are available upon reasonable request to the authors

## Footnotes

1 For example, an organization may choose to implement the recommendation to limit multiple concurrent charts open if that is a criterion on which an accreditation body is evaluating adherence, notwithstanding a lack of evidence in the empirical research to support the practice.

## Notes

### Competing Interest Statement

The authors have declared no competing interest.

### Funding Statement

This study did not receive any funding

